# DALES - a prospective cross-sectional study of incidence of penicillin allergy labels, risk of true allergy and attitudes of patients and anaesthetists to de-labelling strategies

**DOI:** 10.1101/2020.07.02.20144071

**Authors:** L Savic, C Thomas, D Fallaha, Michelle Wilson, PM Hopkins, S Savic, SH Clark, RAFT collaborators (see Supplementary materials)

## Abstract

**Background:** Direct drug provocation testing (DPT) in patients with low-risk penicillin allergy labels would allow population-level ‘de-labelling’. We sought to determine the incidence and nature of penicillin allergy labels in a large UK surgical cohort and to define patient and anaesthetist attitudes towards penicillin allergy testing.

**Methods:** A prospective cross-sectional study was performed in 213 UK hospitals. ‘Penicillin allergic’ patients were interviewed and risk-stratified. Knowledge and attitudes around penicillin allergy were defined in patients and anaesthetists, determining potential barriers to widespread testing.

**Findings:** Of 21,281 patients 12% self-reported penicillin allergy and 67% of these were potentially suitable for direct DPT (stratified low or intermediate risk). Irrespective of risk category 62% wanted allergy testing. Of 4,978 anaesthetists 40% claimed to routinely administer penicillin when they judge the label to be low-risk; 64% would then tell the patient they had received penicillin. Only 47% of all anaesthetists would be happy to administer penicillin to a patient previously de-labelled by an allergy specialist using direct DPT; the commonest reason not to administer penicillin was perceived lack of support from their hospital. On the study days, 13% of low-risk patients requiring penicillin received it, and 6 patients with high-risk labels received it. There were no adverse events in any of this group. However, 1 patient who received an alternative antibiotic suffered suspected anaphylaxis to this.

**Interpretation:** The majority of patients with a penicillin allergy label may be suitable for direct DPT and demand for testing is high among patients. Anaesthetists demonstrate inconsistent, potentially unsafe prescribing in patients labelled as penicillin allergic. More than half of anaesthetists are not reassured by a negative DPT undertaken by a specialist. Significant knowledge gaps may prevent widespread de-labelling being effectively implemented in surgical patients.

**Funding:** The National Institute of Academic Anaesthesia.

## Background

An estimated 2.7 million people in the UK self-report penicillin allergy (1) but the label is incorrect in up to 95% of cases (2). The label is associated with harm, including increased risk of infection with methicillin-resistant *Staphylococcus aureus, Clostridium difficile* and vancomycin-resistant enterococcus, longer hospital stays, and more admissions to critical care (3, 4). A 50% increase in surgical site infections (SSIs) has been demonstrated (5, 6) and in the UK there is also an increased risk of perioperative anaphylaxis attributable to teicoplanin use (7). Most worryingly perhaps, overuse of broad-spectrum antibiotics contributes to the emergence of resistant bacterial strains through selection pressure.

Current penicillin allergy management guidelines in Europe and the US (8-11) recommend a stepwise approach to testing. A detailed history helps delineate true immediate hypersensitivity reactions from side effects; skin testing is then performed to look for evidence of IgE sensitisation. Where negative, the patient undergoes a drug provocation test (DPT) - the gold standard to definitively establish allergic status. Patients who tolerate a DPT are ‘de-labelled’ and can receive penicillins with no risk above that of the baseline population for having a future reaction to penicillin.

This approach is labour-intensive and expensive. In addition, skin testing becomes less reliable over time (12), has significant false positive and negative rates (13), and requires expert interpretation. A critical shortage of trained allergists makes it impossible to provide this expertise for everyone (14). There is growing interest in de-labelling programmes that use risk stratification models to identify patients at low risk of true allergy, who can proceed directly to a DPT without prior skin testing (15-19). In some programmes, risk stratification identifies patients at such low risk of true allergy that the label can be removed with no testing (20). Such programmes are often delivered by non-allergists. There is no universally accepted process for risk stratification, but the model in this study uses many of the key features of those used elsewhere.

There are several potential barriers to widespread de-labelling, and uncertainty about whether de-labelling translates into future penicillin use (21). We sought to describe the scale of self-reported penicillin allergy labels in the UK elective surgical population, to risk stratify patients with the label, and examine attitudes to de-labelling within this population. We also sought to understand anaesthetists’ knowledge and attitudes towards penicillin allergy and de-labelling.

## Methods

A UK-wide cross-sectional observational study was conducted across 213 NHS hospitals. Sites selected 3 data collection days between 21 March^st^ and 31^st^ August 2018. The study comprised a structured interview for patients, a clinician survey for anaesthetists, and a validation survey for sites, detailing local antimicrobial guidelines. For full inclusion and exclusion criteria see Supplementary Material. The study was conducted through the Research and Audit Federation of Trainees (RAFT), a UK-wide network of anaesthetic trainees in collaboration with local research teams (22). The study gained ethics approval (REC reference 17/LO/2106) and HRA approval (IRAS ID 232512).

The STROBE checklist for cross-sectional studies was used to guide reporting of this study. In this paper we present only the results of the study relating to penicillin allergy. The data collected on non-penicillin allergies and other aspects of the anaesthetic survey are to be presented separately.

### 1. Patient Study

Consenting patients provided data on age, gender, history of atopy or urticaria and any drug allergies. Patients reporting penicillin allergy were asked about this in more depth. We included patients who reported either ‘allergy’ or ‘sensitivity’ to penicillin since these terms are used interchangeably. See Supplementary Material for patient survey. Three pictures were shown to participants reporting rash as a symptom, to help increase the reliability of the rash description. These were an urticarial type rash, a maculopapular type rash, and oral thrush.

Patients labelled as penicillin allergic and who required penicillin as first-line choice for antimicrobial prophylaxis were followed up on the day of surgery. The anaesthetic chart was examined by a member of the study team post-operatively for evidence of possible anaphylaxis; specifically, the unplanned use of adrenaline, steroid or antihistamine, mast cell tryptase sampling, unplanned admission to intensive care, or a comment on the chart that anaphylaxis may have occurred.

A process of risk stratification was applied to the penicillin allergy patient histories (see Fig 1). Patients were defined as low risk of allergy when describing side-effects such as nausea, vomiting or thrush, and high-risk if symptoms were suggestive of an immediate type 1 hypersensitivity reaction. Remaining patients, including those who could not remember what had happened or reported ‘other side-effects’, were defined as intermediate risk. Intermediate risk patients reporting uneventful penicillin use since the index event were re-classified as low risk.

Fig 1: Risk stratification model for likelihood of true penicillin allergy

**Fig 1:**
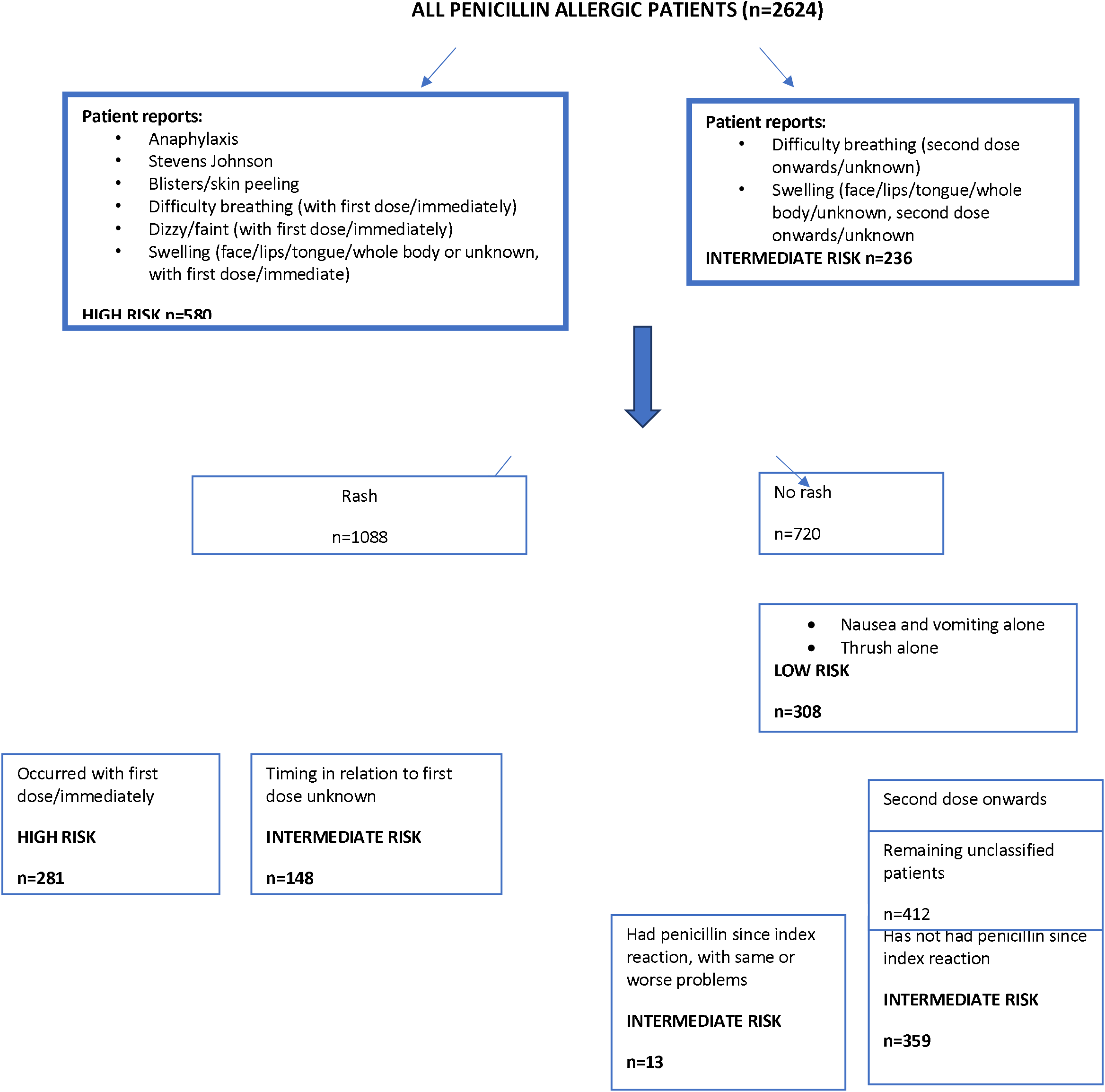
Risk stratification of penicillin allergic patients

### 2. Anaesthetist survey

Data collected included grade of doctor and age range. Knowledge and attitudes were explored using a combination of closed questions and clinical scenarios. Participants were asked what they would prescribe for a patient previously labelled as penicillin allergic but subsequently de-labelled by an allergy specialist using a direct DPT to amoxicillin. See Supplementary Materials for anaesthetist survey. The anaesthetist survey was anonymous; specific anaesthetists were not linked to patients they later anaesthetised.

### 3. Site survey

An additional survey was conducted at each participating site to determine local guidelines for antimicrobial prophylaxis and prescribing in penicillin allergic patients. See Supplementary Materials.

#### Data handling and statistical analysis

Study data were collected and managed using REDCap (Research Electronic Data Capture) hosted at Anaesthesia.Audit on Scotlands Health on the Web secure servers. REDCap is a secure, web-based software platform designed to support data capture for research studies, providing 1) an intuitive interface for validated data capture; 2) audit trails for tracking data manipulation and export procedures; 3) automated export procedures for seamless data downloads to common statistical packages; and 4) procedures for data integration and interoperability with external sources (23, 24). For details on data handling see Supplementary Materials.

Patient characteristics were summarised and differences between pertinent groups (patients with and without penicillin labels, risk stratification groups) were compared using chi-squared tests. Where appropriate, univariable logistic regression was used to assess associations between predictor variables and binary outcome, with multivariable logistic regression used to assess independence of predictors. All statistical analysis was carried out in R, significance tests were all two-sided and p-values < 0.05 were considered significant.

## Results

### 1. Patient Study

A total of 21,281 patients (see Table 1 for patient characteristics) consented to the study. Of these, 2624 (12%) self-reported ‘allergy’ and/or ‘sensitivity’ to penicillin. Among penicillin allergic patients, 274 (10%) also described allergy to at least one other antibiotic and 955 (36%) described allergy to at least one other non-antibiotic drug. In the penicillin allergic group, 68% were female compared with 54% of patients without this label and 56% of all patients. In univariable logistic regression, males were less likely to have a penicillin allergy label than females (OR=0.55, 95%CI: (0.51,0.60), p-value<0.01). There was evidence of an increasing risk of having a penicillin allergy label with increasing age; for example, patients in the 51-75 age group were more likely to report allergy than the 18-25 year group (OR 1.54, 95% CI: (1.20,1.90), p-value <0.01) (Table 2).

**Table 1:** Patient characteristics.

| Characteristic |  | All n=21281 | No PenA label<br>n=18657 | PenA label<br>n=2624 | p-value<br>(from Chi-squared) |
| --- | --- | --- | --- | --- | --- |
| Patient's age range | 18-25 | 1111 (5.2) | 1014 (5.4) | 97 (3.7) | <0.01 |
|  | 26-50 | 6040 (28.4) | 5321 (28.5) | 719 (27.4) |  |
|  | 51-75 | 9879 (46.4) | 8610 (46.1) | 1269 (48.4) |  |
|  | >75 | 4251 (20) | 3712 (19.9) | 539 (20.5) |  |
| Gender | Female | 11939 (56.1) | 10146 (54.4) | 1793 (68.3) | <0.01 |
|  | Male | 9342 (43.9) | 8511 (45.6) | 831 (31.7) |  |
| Type of surgery | Breast | 334 (4.3) | 248 (4.9) | 86 (3.3) | <0.01 |
|  | Cardiac | 75 (1) | 45 (0.9) | 30 (1.1) |  |
|  | Chronic Pain | 227 (3) | 155 (3.1) | 72 (2.7) |  |
|  | Colorectal | 242 (3.1) | 165 (3.3) | 77 (2.9) |  |
|  | Dental | 153 (2) | 90 (1.8) | 63 (2.4) |  |
|  | ENT / Head & Neck | 633 (8.2) | 406 (8) | 227 (8.7) |  |
|  | General surgery (including UGI / HPB) | 702 (9.1) | 491 (9.7) | 211 (8) |  |
|  | Gynaecology | 893 (11.6) | 574 (11.3) | 319 (12.2) |  |
|  | Neuro | 107 (1.4) | 80 (1.6) | 27 (1) |  |
|  | Non-theatre | 66 (0.9) | 42 (0.8) | 24 (0.9) |  |
|  | Obstetrics | 199 (2.6) | 127 (2.5) | 72 (2.7) |  |
|  | Ophthalmology | 1102 (14.3) | 699 (13.8) | 403 (15.4) |  |
|  | Orthopaedics | 1527 (19.9) | 1040 (20.5) | 487 (18.6) |  |
|  | Plastics | 284 (3.7) | 183 (3.6) | 101 (3.8) |  |
|  | Spinal | 104 (1.4) | 66 (1.3) | 38 (1.4) |  |
|  | Thoracics | 57 (0.7) | 41 (0.8) | 16 (0.6) |  |
|  | Transplant | 7 (0.1) | 6 (0.1) | 1 (0) |  |
|  | Urology | 801 (10.4) | 498 (9.8) | 303 (11.5) |  |
|  | Vascular | 176 (2.3) | 109 (2.2) | 67 (2.6) |  |
|  | Not recorded | 13592 (-) | 13592 (-) | 0 (-) |  |

**Table 2:**
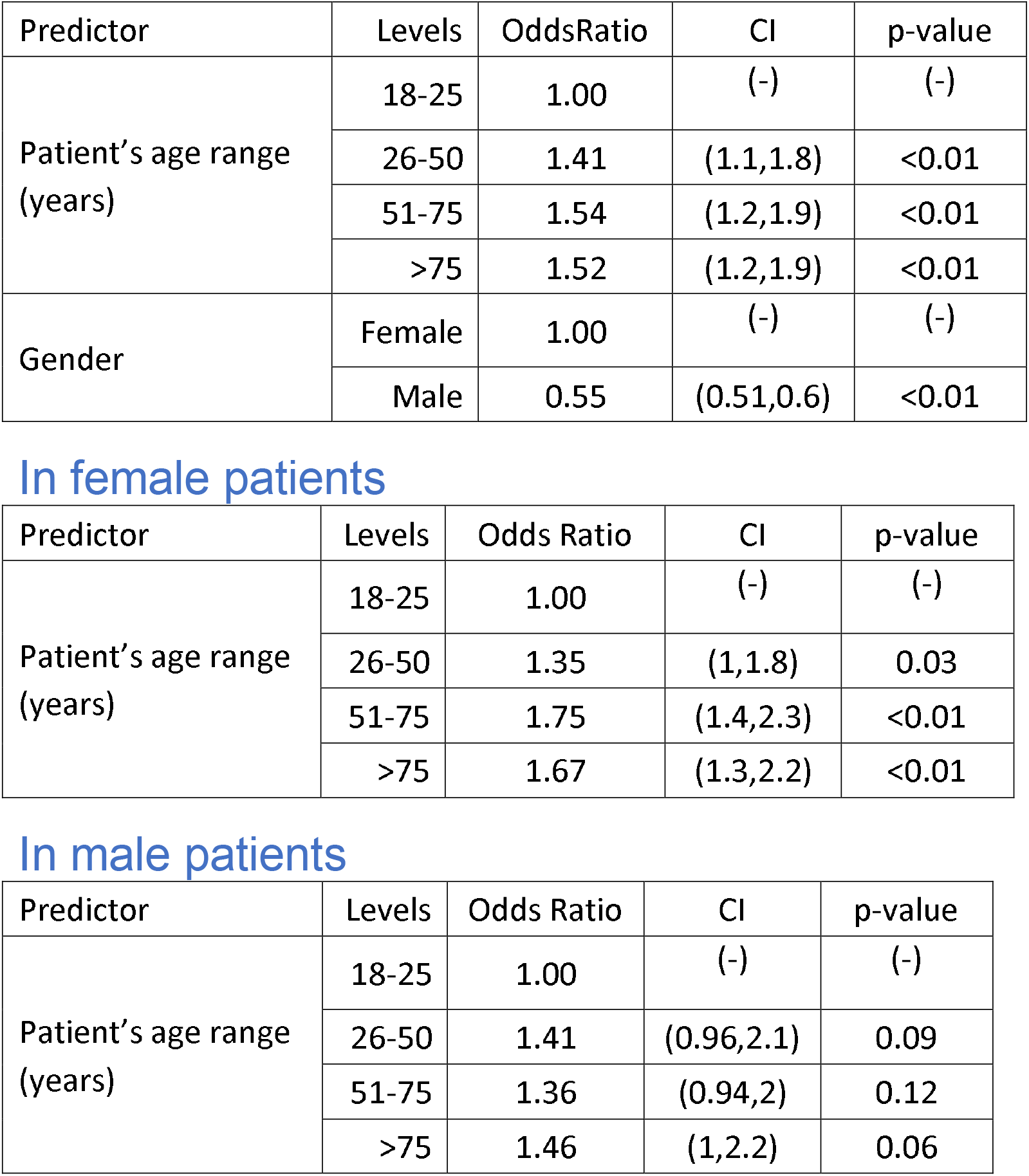
Logistic regression with ‘penicillin allergic’ status as dependent variable In all patients.

| Predictor | Levels | OddsRatio | CI | p-value |
| --- | --- | --- | --- | --- |
| Patient's age range (years) | 18-25 | 1.00 | (-) | (-) |
|  | 26-50 | 1.41 | (1.1,1.8) | <0.01 |
|  | 51-75 | 1.54 | (1.2,1.9) | <0.01 |
|  | >75 | 1.52 | (1.2,1.9) | <0.01 |
| Gender | Female | 1.00 | (-) | (-) |
|  | Male | 0.55 | (0.51,0.6) | <0.01 |

**In female patients**
| Predictor | Levels | Odds Ratio | CI | p-value |
| --- | --- | --- | --- | --- |
| Patient's age range (years) | 18-25 | 1.00 | (-) | (-) |
|  | 26-50 | 1.35 | (1,1.8) | 0.03 |
|  | 51-75 | 1.75 | (1.4,2.3) | <0.01 |
|  | >75 | 1.67 | (1.3,2.2) | <0.01 |

**In male patients**
| Predictor | Levels | Odds Ratio | CI | p-value |
| --- | --- | --- | --- | --- |
| Patient's age range (years) | 18-25 | 1.00 | (-) | (-) |
|  | 26-50 | 1.41 | (0.96,2.1) | 0.09 |
|  | 51-75 | 1.36 | (0.94,2) | 0.12 |
|  | >75 | 1.46 | (1,2.2) | 0.06 |

Rash was the most commonly reported feature (n=1445, 55%), and occurred as a sole sign in 63% of those who reported it. Of patients with a rash, 795 (55%) stated it resembled a maculo-papular rash in appearance (picture B in Appendix 3). See Table 3 for all symptoms reported.

**Table 3:** Nature of penicillin allergy: absolute numbers for each symptom, and relative incidence of each symptom.

| Characteristic | Level | All n=2624 | Patients with only each symptom* |
| --- | --- | --- | --- |
| Rash | Yes | 1446 (55.1) | 912 (63.1) |
|  | No | 1178 (44.9) | 543 (36.9) |
| Blisters / skin peeling | Yes | 173 (6.6) | 47 (27.2) |
|  | No | 2451 (93.4) | 126 (72.8) |
| Difficult to breathe and / or became wheezy | Yes | 219 (8.3) | 21 (9.6) |
|  | No | 2405 (91.7) | 198 (90.4) |
| Swelling | Yes | 529 (20.2) | 172 (32.5) |
|  | No | 2095 (79.8) | 357 (67.5) |
| Dizzy or faint | Yes | 146 (5.6) | 33 (22.6) |
|  | No | 2478 (94.4) | 113 (77.4) |
| Sick / vomited / had a sore stomach / had diarrhoea | Yes | 482 (18.4) | 241 (50) |
|  | No | 2142 (81.6) | 241 (50) |
| Thrush | Yes | 55 (2.1) | 33 (60) |
|  | No | 2569 (97.9) | 22 (40) |
| Anaphylaxis or a serious reaction | Yes | 131 (5) | 64 (48.9) |
|  | No | 2493 (95) | 67 (51.1) |
| Stevens Johnson syndrome / DRESS **/ AGEP*** | Yes | 1 (0) | 0 (0) |
|  | No | 2623 (100) | 2624 (100) |
| Other side effect | Yes | 175 (6.7) | 95 (54.3) |
|  | No | 2449 (93.3) | 80 (45.7) |
| Unknown | Yes | 304 (11.6) | 289 (95.1) |
|  | No | 2320 (88.4) | 15 (4.9) |

\*patients with only symptom summarized in column 1 e.g. 1446 people had a rash and of these 1446-912 only had a rash and 543 also experienced another symptom.
\*\*Drug Reaction with Eosinophilia and Systemic Symptoms
\*\*\*Acute Generalised Exanthematous Pustulosis

Using the stratification model (Fig. 1), we determined that 27% (n=715) reported low-risk histories, 33% (n=861) high-risk histories, and 40% (n=1048) intermediate-risk histories. There was a greater likelihood of women having a high-risk label than men (OR for men 0.81, CI 0.68-0.97, p=0.02).

Since the utility of skin testing to risk stratify patients decreases significantly over time, we sought to determine the proportion of historic reactions (>10 years) in our cohort. In the low risk group 68% reported the index reaction as being > 10 years, 88% in intermediate-risk, and 73% in the high-risk group. Since the intermediate group had no features consistent with genuine allergy, all those with low or intermediate risk might together be considered suitable for direct DPT, representing 67% (95% CI 65.2 – 68.8%) of all patients labelled as penicillin allergic in this population.

A minority of all patients reporting penicillin allergy (n=141, 5.4%) recalled having had undergone previous allergy testing, although the nature of this testing (skin testing alone, skin testing plus DPT, or DPT alone) was not elucidated Most recalled a positive result (n=95, 67%) but 25% (n=35) could not remember the result. In those with positive test results, the majority (58, 61%) had a high-risk index reaction history. Of all those who had not previously received testing, 62%, (n=1541) stated that they would like to be tested. Among those not wanting testing (n=940, 38%), the single most common reason was “I would never take penicillin again, whatever the result” (n=408, 43%). See Supplementary Materials. The risk category of the patient did not appear to influence the likelihood of wanting to be tested (58% low-risk vs 62% intermediate-risk vs 55% of high-risk patients). Multivariable analysis demonstrated an effect of age on whether the patient wishes to be tested, with those in the >75 years age range less likely to want to undergo testing (OR 0.34, CI 0.2-0.55, p-value <0.01). There was also an association with other patient characteristics; those reporting atopy were more likely to want testing (OR 1.21, CI 1.01-1.45, p-value = 0.03), and those reporting sickness as their presenting ‘allergic’ feature were less likely to (OR 0.64, CI 0.51-0.81, p-value <0.01). Among those patients who wanted testing (n=1541), the majority (68%, n=1060) would be happy to have the label removed by an allergy specialist on the basis of history alone.

In the group with a penicillin allergy label, 526 (20%) were listed for surgery requiring penicillin as first-line antimicrobial prophylaxis choice. Penicillin was administered despite the allergy label in 33 of these patients (6%), of whom 6 had a high-risk history, 8 an intermediate history, and 19 a low-risk history. Second-line prophylaxis was given to 52% of the penicillin allergic patients (n=251), while 39% (n=203) were given no antibiotic at all or an antibiotic which was non-standard in that hospital. Included in this latter group were an unidentified number of patients for whom antibiotic use was contingent on intraoperative events and may not have been required (eg. antibiotic use in laparoscopic cholecystectomy only if the bile duct is injured). Two patients who received alternative antibiotics suffered an adverse event in keeping with anaphylaxis during surgery (see Supplementary Materials). In one, anaphylaxis to a glycopeptide antibiotic was later confirmed at allergy clinic. No further details are available for the second patient. None of the patients who received penicillin had an adverse intraoperative event.

### 2. Anaesthetist Study

A total of 4978 anaesthetists participated of whom 64% (n=3051) were consultant grade, 12% associate specialists or staff grades, and the remainder junior grade doctors (n=1158, 24%) or physician assistants (n=23, 0.5%). There was mixed understanding of which symptoms/signs were likely to reflect true allergy versus side-effect, with 5% (n=214) stating that a history of ‘anaphylaxis’ was unlikely or highly unlikely to represent true allergy; 7% (n=322) stating that nausea/diarrhoea was likely or highly likely to represent true allergy; and 4% (n=177) stating that a patient with a history of penicillin allergy who had subsequently received a penicillin without adverse effect was still likely /highly likely to have a true allergy. The majority of anaesthetists (n=3158, 66%) thought that a true allergic reaction was still likely/highly likely if the symptoms only developed after taking the second or subsequent doses. See Supplementary Materials.

When prescribing antibiotics to patients with penicillin allergy labels, 40% (n=1934) of anaesthetists stated they would give penicillin if they felt the label was ‘highly unlikely to represent true allergy’, and 60% (n=2829) stated they would always avoid penicillin in this situation. In the group happy to overrule the allergy label based on their own judgment we asked what actions they would take following uneventful administration of penicillin, to inform other healthcare providers or the patient. The majority (n=1357, 70%) would amend the anaesthetic chart, but few other reported actions would consistently be taken, with 14% (n=266) telling the general practitioner and 63% telling the patient. See Supplementary materials. Among anaesthetists who would always avoid penicillin in patients with a penicillin allergy label, there were multiple reasons for this behaviour with perceived lack of support from the hospital the single most common (Fig 2).

**Fig 2.**
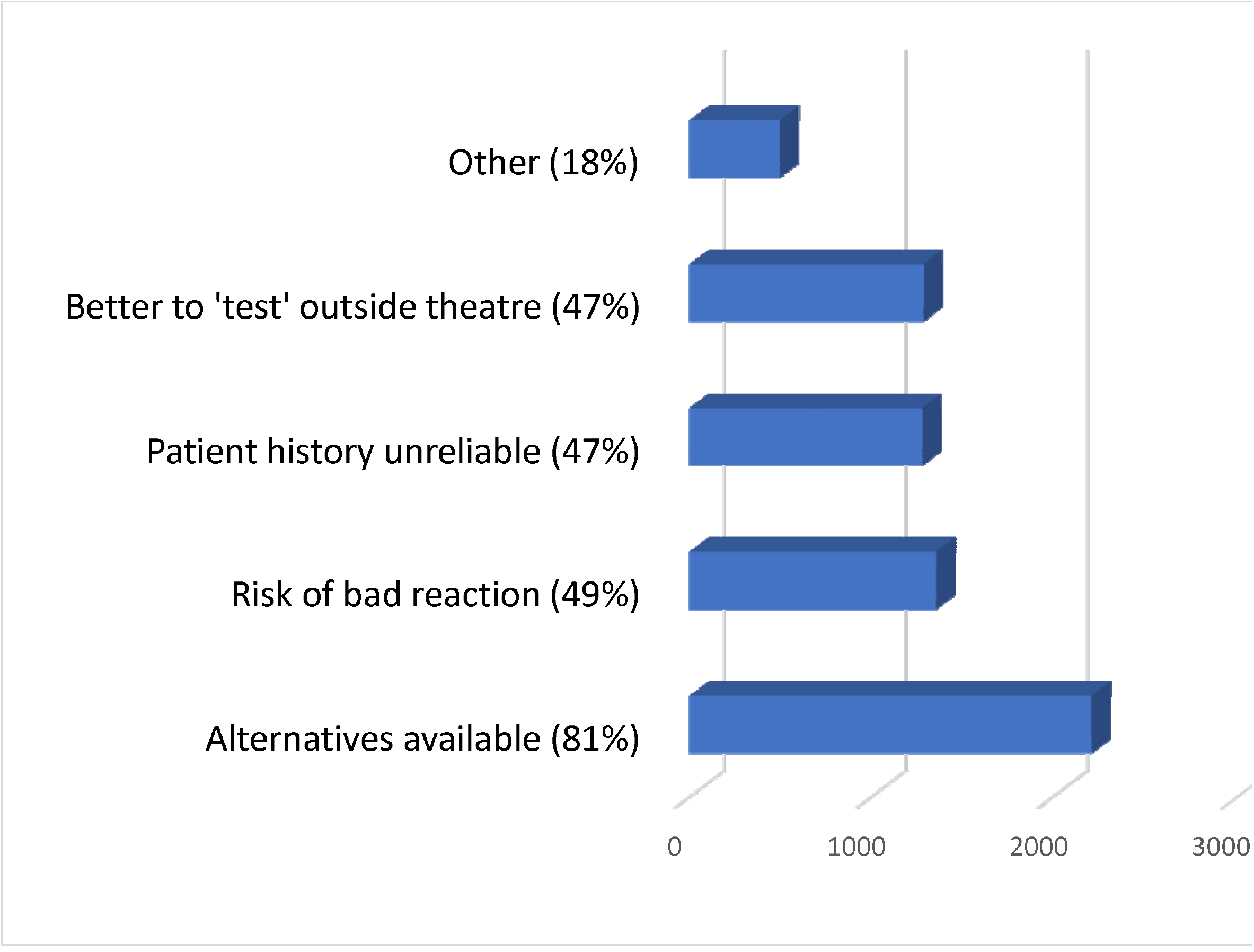
Why do 60% of anaesthetists “always avoid” giving penicillin?

Anaesthetists were asked whether they would administer a penicillin to a patient who had previously been de-labelled by an allergy specialist following an uneventful direct DPT with amoxicillin. Just under half (47%, n=2240) stated that they would administer a penicillin, with the remainder responding ‘no’ (n=633, 13%) or ‘unsure’ (n=1828, 38%). The two most common reasons were that they would require formal guidelines from their hospital to support this action (n=1379, 56%), and that patients should undergo skin testing in order to be de-labelled (n=1247, 51%). There was concern that an oral DPT was an insufficient test for patient requiring intravenous penicillin (n=667, 27%). See Table 4. Of those anaesthetists confident to give penicillin to a patient whose label was ‘highly unlikely’ to be correct, only around half (n=1038, 54%) would be happy to give penicillin to a patient de-labelled using direct DPT by a specialist. Among anaesthetists not happy to give penicillin to ‘highly unlikely’ labels, 42% (n=1193) would be happy to give penicillin if the patient was de-labelled by a specialist. Among those anaesthetists who would accept the results of an oral DPT the majority (n=1856, 83%) stated that if the allergy specialist had ‘de-labelled’ the patient on the basis of history alone (no skin testing or DPT) they would still be confident to administer a penicillin in theatre. Where anaesthetists would not accept de-labelling by a specialist on the basis of history alone, this was most commonly because they felt testing of some sort was essential to de-labelling.

**Table 4:** In anaesthetists who stated ‘no’ or ‘unsure’ to whether they would give penicillin to a patient who has been de-labelled by a specialist using a direct oral DPT, reasons for not doing so.

| Reason |  | No<br>n=633 | Unsure<br>n=1828 |
| --- | --- | --- | --- |
| My understanding is that patients should also be skin tested | Yes | 372<br>(58.8) | 875 (47.9) |
|  | No | 261<br>(41.2) | 953 (52.1) |
| The penicillin received during testing might not be the same one I give in theatre | Yes | 162<br>(25.6) | 362 (19.8) |
|  | No | 471<br>(74.4) | 1466 (80.2) |
| I will be giving intravenous penicillin during surgery but the testing was oral | Yes | 201<br>(31.8) | 466 (25.5) |
|  | No | 432<br>(68.2) | 1362 (74.5) |
| I would never give penicillin to someone previously labelled as allergic, whatever the result of testing | Yes | 70<br>(11.1) | 57 (3.1) |
|  | No | 563<br>(88.9) | 1771 (96.9) |
| I would require clear local guidelines to support the use of penicillin in this situation | Yes | 298<br>(47.1) | 1081 (59.1) |
|  | No | 335<br>(52.9) | 747 (40.9) |
| Other reason(s) | Yes | 35 (5.5) | 145 (7.9) |
|  | No | 598<br>(94.5) | 1683 (92.1) |

*Multiple selections allowed*

When asked about the use of test doses for antibiotics, 49% (2319) stated they ‘never’ gave them, the remainder giving test doses routinely, or in selected patients. Anaesthetists who avoid penicillin in anyone with the penicillin allergy label were more likely to give a test dose routinely than anaesthetists who are happy to administer penicillin to someone with a label they judged to be incorrect (31% vs 24%). See Supplementary Materials.

Anaesthetists were not always aware of whether their site had guidelines on prescribing in patients with a penicillin allergy label. Of 203 sites, 63 had specific guidelines for how to assess risk of true penicillin allergy in patients with the label and prescribe accordingly. and Expressed as the median range percentage, anaesthetists thought that such guidance existed in 30%. Conversely, where guidance did not exist, anaesthetists thought it did in 27% of sites. Among anaesthetists 52% (n=2474) would avoid cephalosporins in patients with a label of penicillin allergy; 17% (n=804) would routinely prescribe a cephalosporin, and a further 25% (n=1154) would follow local guidelines on cephalosporin use. Of 203 sites, 97 had guidelines on prescribing of cephalosporins to patients with the label.

## Discussion

This is the largest prospective study to date examining incidence of penicillin allergy in an unselected elective surgical population and the first to prospectively perform detailed risk stratification in these patients. We have demonstrated a high incidence of reported penicillin allergy in this population and a greater likelihood of older, female patients having the label. Up to 67% of patients with the allergy label appear suitable to undergo direct DPT; this approach significantly reduces the time and cost burden of testing and might allow widespread testing in this population. It is important to note that this is a population-based study and that all patients with the label require individualised assessment before testing.

Our study defines some key attitudes among patients. A sub-group state they would never take penicillin again irrespective of test result; these patients may benefit from education about the potential harms of avoiding penicillin. Overall, however, there was high demand for allergy testing and this was largely unaffected by the severity of the index reaction. Patients do not appear to have preconceived ideas about how testing should be performed, with a majority stating that if an allergy specialist judged their reaction to be non-allergic on the basis of history alone, they would be happy to be de-labelled without formal testing.

We have determined for the first time some key attitudes, knowledge and behaviours among anaesthetists in relation to penicillin allergic patients. A minority were unable to appropriately categorise allergy histories which were clearly low- or high-risk, and there was widespread misunderstanding around the significance of symptom onset in relation to the first dose administered. These discrepancies may simply reflect misinterpretation of the questions however (e.g. ‘after the second (lifetime) exposure to penicillin’, rather than ‘after the second dose of an individual course), or simple error in using the 0-5 scale.

We have demonstrated mixed prescribing habits in penicillin allergic patients with up to 40% of anaesthetists stating that they would administer penicillin to a patient with the label they judged to be incorrect. In the absence of training or specialist drug allergy knowledge this represents a potential patient safety issue. We found that anaesthetists, having given penicillin uneventfully, would not then cascade this information to other healthcare professionals or the patient, negating any long-term benefits from de-labelling. This also raises the issue of whether patients are appropriately consented for what is in effect a DPT. There is an apparent discrepancy between what anaesthetists say they will do and what they actually do; with 40% claiming they would prescribe penicillin to low-risk label patients, but only 13% of low-risk patients who required penicillin on the study days receiving it.

However, the most significant finding is that fewer than half of anaesthetists would be confident administering penicillin to a patient who has previously been de-labelled by an allergy specialist using direct DPT. Key reasons for this include misunderstanding of allergy testing and perceived lack of support from their hospital. These are likely to be the greatest barrier to any effective programme of systematic de-labelling in surgical patients and could potentially be addressed with greater education and structured guidance within hospitals.

Other findings of note include the high incidence of penicillin allergic patients receiving non-standard antibiotics for surgery, or no antibiotics at all. We cannot determine from our data what the reasons were for this, who made the antibiotic decisions within the theatre team, or what contribution this might make to the well described poor outcomes in surgical patients with the label.

There are several limitations to this study. Firstly, the data collected on symptoms of the index reaction were limited; with greater detail, risk stratification may have changed. Secondly, the inclusion of pictures to help patients to describe the rash they experienced may not have added value to the description and may have been misleading. The quality of the pictures shown is likely to have varied depending on the electronic device used to display these. Thirdly, since the anaesthetic survey was anonymous we did not link individual anaesthetists to their patients and therefore could not identify differences between stated and actual prescribing habits on the study days for any individual. Lastly, as we did not collect longer term follow-up data on outcomes in patients who required penicillin during surgery, we are unable to comment on incidence of infection, length of stay and readmission rates.

## Conclusion

Penicillin allergy labels are easy to acquire and difficult to lose. We have identified some key attitudes and behaviours among both doctors and patients which might be relevant to this problem. Our findings are likely to be representative of the UK elective surgical population because of sample size and may translate across different groups of patients and medical specialities.

We demonstrate high demand for testing among patients and that between 25-67% might be suitable for direct DPT. Anaesthetists exhibit contradictory and potentially unsafe prescribing habits in patients with the label and there are several important misconceptions around penicillin allergy testing. The persistent avoidance of penicillin by clinicians in the face of negative testing is a key problem and warrants further exploration; de-labelling is futile if it doesn’t translate to future penicillin use. Some concerns might be allayed with additional guidance from hospitals. However, if anaesthetists are to take ownership of the problem of incorrect penicillin allergy labels as part of perioperative medicine, there is also a significant educational gap to bridge.

## Data Availability

Supplementary materials will be available online on request

## Acknowledgements

More than 1500 anaesthetic trainees, consultants, and research nurses collected data for this study on behalf of RAFT and we are extremely grateful for all their hard work and commitment. We also thank the National Institute of Academic Anaesthesia for funding this work, and Leeds Teaching Hospitals Trust who were the sponsor.

## References

1. West RM, Smith, C.J, Pavitt, S.H, et al. “Warning: allergic to penicillin:” Association between penicillin allergy status in 2.3 million NHS general practice electronic health records, antibiotic prescribing, and health outcomes. J Antimicrob Chemotherapy. 2019.

2. Mohamed OE, Beck S, Huissoon A, Melchior C, Heslegrave J, Baretto R, et al. A Retrospective Critical Analysis and Risk Stratification of Penicillin Allergy De-labelling in a UK Specialist Regional Allergy Service. J Allergy Clin Immunol Pract. 2018.

3. Blumenthal KG, Lu N, Zhang Y, Li Y, Walensky RP, Choi HK. Risk of meticillin resistant Staphylococcus aureus and Clostridium difficile in patients with a documented penicillin allergy: population based matched cohort study. BMJ. 2018;361:k2400.

4. Charneski L, Deshpande G, Smith SW. Impact of an antimicrobial allergy label in the medical record on clinical outcomes in hospitalized patients. Pharmacotherapy. 2011;31(8):742–7.

5. Blumenthal KG, Ryan EE, Li Y, Lee H, Kuhlen JL, Shenoy ES. The Impact of a Reported Penicillin Allergy on Surgical Site Infection Risk. Clin Infect Dis. 2018;66(3):329–36.

6. Pool C, Kass J, Spivack J, Nahumi N, Khan M, Babus L, et al. Increased Surgical Site Infection Rates following Clindamycin Use in Head and Neck Free Tissue Transfer. Otolaryngol Head Neck Surg. 2016;154(2):272–8.

7. harper NJN CT, Garcez T, Farmer L, Floss K, Marinho S, Torevell H, Waner A, Ferguson K, Hitchman J, Egner W, Kemp H, Thomas M, Lucas DN, Nasser S, Karanam S, Kong K-L, Farooque S, Bellamy M, McGuire N. Anaesthesia, surgery, and life-threatening allergic reactions: epidemiology and clinical features of perioperative anaphylaxis in the 6th National Audit Project (NAP6). British Journal of Anaesthesia. 2018.

8. Mirakian R, Ewan PW, Durham SR, Youlten LJ, Dugue P, Friedmann PS, et al. BSACI guidelines for the management of drug allergy. Clin Exp Allergy. 2009;39(1):43–61.

9. Bernstein IL, Li JT, Bernstein DI, Hamilton R, Spector SL, Tan R, et al. Allergy diagnostic testing: an updated practice parameter. Ann Allergy Asthma Immunol. 2008;100(3 Suppl 3):S1–148.

10. Demoly P, Adkinson NF, Brockow K, Castells M, Chiriac AM, Greenberger PA, et al. International Consensus on drug allergy. Allergy. 2014;69(4):420–37.

11. Garvey LH. The use of drug provocation testing in the investigation of suspected immediate peri-operative allergic reactions: current status. Br J Anaestesia. 2019;123:e:126-34.

12. Blanca M, Torres MJ, Garcia JJ, Romano A, Mayorga C, de Ramon E, et al. Natural evolution of skin test sensitivity in patients allergic to beta-lactam antibiotics. J Allergy Clin Immunol. 1999;103(5 Pt 1):918–24.

13. Banks TA, Tucker M, Macy E. Evaluating Penicillin Allergies Without Skin Testing. Curr Allergy Asthma Rep. 2019;19(5):27.

14. NICE. Drug allergy: diagnosis and management. https://www.nice.org.uk/guidance/cg183/chapter/Introduction. 2014.

15. du Plessis T, Walls G, Jordan A, Holland DJ. Implementation of a pharmacist-led penicillin allergy de-labelling service in a public hospital. J Antimicrob Chemother. 2019.

16. Savic LG, L. Kaura, V. Toolan, J. Sandoe, J. A.T. Hopkins, P.M. SavicS., Penicillin Allergy De-Labelling Ahead of Elective Surgery – Is it Feasible and What are the Barriers? British Journal of Anaesthesia. 2018.

17. Tucker MH, Lomas CM, Ramchandar N, Waldram JD. Amoxicillin challenge without penicillin skin testing in evaluation of penicillin allergy in a cohort of Marine recruits. J Allergy Clin Immunol Pract. 2017;5(3):813–5.

18. Confino-Cohen R, Rosman Y, Meir-Shafrir K, Stauber T, Lachover-Roth I, Hershko A, et al. Oral Challenge without Skin Testing Safely Excludes Clinically Significant Delayed-Onset Penicillin Hypersensitivity. J Allergy Clin Immunol Pract. 2017;5(3):669–75.

19. Iammatteo M, Alvarez Arango S, Ferastraoaru D, Akbar N, Lee AY, Cohen HW, et al. Safety and Outcomes of Oral Graded Challenges to Amoxicillin without Prior Skin Testing. J Allergy Clin Immunol Pract. 2018.

20. Blumenthal KG, Wolfson, A.R., Hsu, J.T. Outcomes of beta-lactam antibiotic test dose procedures for patients with reported beta-lactam allergy performed in a large US healthcare system. J of Allergy and CLinic Immunol: In Practice. 2019;143(2):AB25.

21. Rimawi RH, Shah KB, Cook PP. Risk of redocumenting penicillin allergy in a cohort of patients with negative penicillin skin tests. J Hosp Med. 2013;8(11):615–8.

22. Trainees RaAFo. 2020.

23. PA Harris RT, BL Minor, V Elliott, M Fernandez, L O’Neal, L McLeod, G Delacqua, F Delacqua, J Kirby, SN Duda, REDCap Consortium. The REDCap consortium: Building an international community of software partners,. J Biomed Inform. 2019.

24. PA Harris RT, R Thielke, J Payne, N Gonzalez, JG. Conde. Research electronic data capture (REDCap) – A metadata-driven methodology and workflow process for providing translational research informatics support,. J Biomed Inform. 2009;42(2):377–81.

